# COVID-19 Growth Rate Decreases with Social Capital

**DOI:** 10.1101/2020.04.23.20077321

**Authors:** Lav R. Varshney, Richard Socher

## Abstract

**Background:** Social capital has been associated with many public health variables including mortality, obesity, diabetes, and sexually-transmitted disease rates. However, the relationship of social capital to the spread of infectious disease like COVID-19 is lacking. The COVID-19 pandemic presents an unprecedented threat to global health and economy, for which control strategies have relied on aggressive social distancing. However, an understanding of how social capital is related to changes in human mobility patterns for adherence to social distancing is lacking.

**Objective:** This study examines the association between state- and county-level social capital indices and community health indices in the United States, and the growth rate of COVID-19 cases. It also examines changes in human mobility.

**Methods:** Using publicly available state- and county-specific time series data for COVID-19 cases from March 13 to March 31, we used exponential fits to determine growth rate. We obtained publicly available mobility change data, originally measured from GPS-enabled mobile devices. The design was then state- and county-level correlation analysis with social capital and community health indices from the Social Capital Project (United States Senate).

**Results:** In bivariate linear correlation analyses, we find social capital and community health indices were negatively associated with COVID-19 growth rates at both the state and county levels. The correlation was strongest at the county level for the community health index: a one-unit increase in the county community health index was associated with a decrease in the COVID-19 growth rate exponent by 0.045. In further bivariate correlation analyses, we find that social capital indices were negatively associated with retail/recreation movement and positively associated with residential movement. That is, an increase in social capital is correlated with slower COVID-19 infection spread and more adherence to social distancing protocols.

**Conclusion:** Our results indicate the potential benefit of incorporating social capital concepts in planning policies to control the spread of COVID-19, e.g. different social distancing requirements in different communities. The results also indicate a need for further research into this potentially causal relationship, including examining interventions to increase social capital, community health, and institutional health.

## 1 Introduction

The concept of social capital was initially developed in the social sciences [1], but has become central to understanding the social determinants of public health over the last two decades. Within public health research, social capital is defined as the resources available to individuals via their membership and active participation in community networks and is said to reside in the structure and quality of relationships among community members [2–4]. Proxy indicators of area-level social capital are typically used to explain variations in health status across geographic localities. For the United States, such social capital indices are available at the state- and county-levels [5, 6]. Metastudies demonstrate the widespread positive impact social capital has on (or associations with) numerous public health variables including mortality, obesity, diabetes, and sexually-transmitted infection rates [7]. There has been little characterization, however, of associations between social capital and other infectious diseases [8,9], including the novel coronavirus disease COVID-19, for which the absence of a vaccine necessitates the use of aggressive social distancing to control its unprecedented global pandemic. Moreover, there has been little characterization of association between social capital and the rate at which movement is decreased under social distancing.

Whereas social capital may build the social infrastructure in a community for individuals to sacrifice for the greater good and self-isolate in response to an infectious disease outbreak, higher levels of trusting social interactions may also increase opportunities for face-to-face disease transmission and accelerate contagion [9, 10]. This study examines the balance between these two possibilities in the case of COVID-19 in the United States, performing an exploratory analysis of state- and county-level relationships between social capital indices (including community health sub-indices) and COVID-19 case growth rate exponents that characterize how quickly the disease is spreading, cf. [11]. The study further considers relationships between social capital indices (including institutional health sub-indices) and actions to reduce movement in response to COVID-19, cf. [12, 13].

Past studies of infectious disease have shown effects both ways. More diverse social networks were associated with greater resistance to upper respiratory illness [14], whereas social participation increased risk of person-to-person spread of influenza among unimmunized older adults [15]. State-level social capital indices in the United States are strongly associated with reduced rates of tuberculosis [9]; a similar but weaker association is found at the neighborhood level for risk of bacterial infections among infants [16]. Considering a mosquito-borne disease rather than one with person-to-person transmission, Indonesian villages with stronger social capital have reduced dengue fever by taking collective action to improve hygiene, manage waste properly, and eradicate mosquito breeding grounds [17].

Independent of the free-rider problem of few people complying with social distancing and thereby accelerating contagion [10], several other causal mechanisms have been put forth to explain the link between social capital and health. These mechanisms for social capital influence include: social isolation linked to poor health, social norms supporting healthy behavior, access to healthcare services, neighbors taking more responsibility for one another, and development of public policies that protect all citizens [9]. We do not address causal mechanisms in this work, but our results indicate a need for further research into the potentially causal relationship between social capital and COVID-19 spread, which may provide more nuance to public health policy.

## 2 Materials and Methods

We consider state- and county-level COVID-19 cases reported in the United States between March 13 and March 31, 2020, taken from The New York Times database (https://github.com/nytimes/covid-19-data). We do not consider more recent case reports beyond March 31 (but see Fig. 8)—which is two weeks (COVID- 19 incubation period) after the first widespread shelter-in-place order was enacted in the country (in the San Francisco Bay area, on March 16)—so we can examine disease spread in the absence of mandatory lockdown orders. Following [18], we exclude areas that had no COVID-19 cases on March 13 in our analysis, whose low case counts are likely due to introductions from outside, and for which accurate estimates of local case growth rate is unlikely. At the state level, this only excludes West Virginia. At the county level, we end up with 311 counties from around the country, which is roughly 10% of all counties. As a side note, counties that had reported COVID-19 cases by March 13 generally have lower social capital and community health indices, see Fig. 6 for a comparison of cumulative distribution functions.

To characterize the spread of COVID-19 in these areas, we assume an exponential growth of cases (valid for this novel virus during the time period we consider since no one yet had immunity) and characterize the growth exponent. In particular, following the method of [18], we subtract deaths from cases in each locality and find the slope of the log-log plot of cases as a function of time for each resulting time series of active COVID-19 cases. As noted there, estimating the growth rate of epidemics accurately is difficult, but we are only concerned with comparing growth rates among localities rather than obtaining precise growth exponent estimates.

These estimated growth exponents are then used as part of correlation analysis. The state- and county-level indices of social capital are obtained from the Social Capital Project of the United States Senate [6]. An alternative county-level social capital metric from Pennsylvania State University is also used for a robustness check [5]. The social capital index from the Social Capital Project is based on sub-indices that measure family unity, family interaction, social support, community health, institutional health, collective efficacy, and philanthropic health. Given our research focus, we also specifically consider the community health index which itself is made up of measures of non-profit organizations, religious organizations, and participation rates in meetings, demonstrations, volunteer work, and helping neighbors. We also specifically consider the institutional health index which itself is made up of measures of voting, census response, and confidence in institutions.

The bivariate relationship between each social capital or community health index and COVID-19 growth rate exponent are assessed using linear regression. The *p*-value of the slope term, the overall *R*^2^ value, and the fitted slope coefficient are all used. Likewise for mobility.

Since there is variability in testing for COVID-19 across geographies, one might wonder if a greater number of reported cases is due to more widespread testing. Due to data availability, we focus on state-level analysis and measure the association between social capital or community health indices and the number of tests performed per capita, cf. [19]. We specifically look at the cumulative total number of tests performed up to March 31, 2020, the end of our cases data period. These test counts are from the COVID Tracking Project (https://covidtracking.com/data). Population data for the states are the July 2019 estimate from the U.S. Census.

To consider an association between social distancing and social capital, we use a mobility index derived from cell phone location data as a coarse measure of distance [20]. In particular, we use the county-level mobility index computed by the firm Cuebiq (https://clara-beta.cuebiq.com/). For individuals, they calculate distance traveled by measuring a line between opposite corners of a box drawn around the locations observed for each person on each day. The travel for each county is the median of these per-person distances. The weekly average is the arithmetic mean of this quantity, and the quantity we consider is the percentage change as compared to the yearly average. We use the week of March 23. This is a fairly coarse measure of mobility and does not capture the kind of fine-grained information about social distancing that contact tracing [21] would. For example, if one’s long commute was replaced by greater social interaction with neighbors, the mobility index would decrease significantly yet social contacts would increase and reduce the effectiveness of contagion control via social distancing.

As a more fine-grained mobility index, we draw on data from Google Community Mobility Reports (https://www.google.com/covid19/mobility/). These report change in movement by state and county, across different categories of places such as retail and recreation, and residential—the two categories we focus on, as they are where one would expect strong decreases and increases, respectively, under social distancing. For each day, the reports provide changes as compared to a baseline value for that day of the week during the 5-week period between January 3 and February 6, 2020. We consider the county level and average the daily percentages for the seven-day period between March 23 and March 29. The data does not include all counties, and so we end up with 2500 counties in our dataset.1

## 3 Results

Figures 1 and 2 show the results of our bivariate analysis at the state and county levels, respectively for both social capital and community health indices. At the state level, we find a statistically significant negative correlation between fitted COVID-19 growth rate exponent and both social capital and community health indices: the more these indices, the slower the COVID-19 spread. The social capital index explains 9.2% of the variation in exponent, and the community health sub-index explains 11.7% of the variation. The effect size is such that each point of increase in social capital index corresponds to a 0.015 decrease in the growth exponent; each point of increase in the community health index corresponds to a 0.017 decrease in growth exponent.

**Figure 1:**
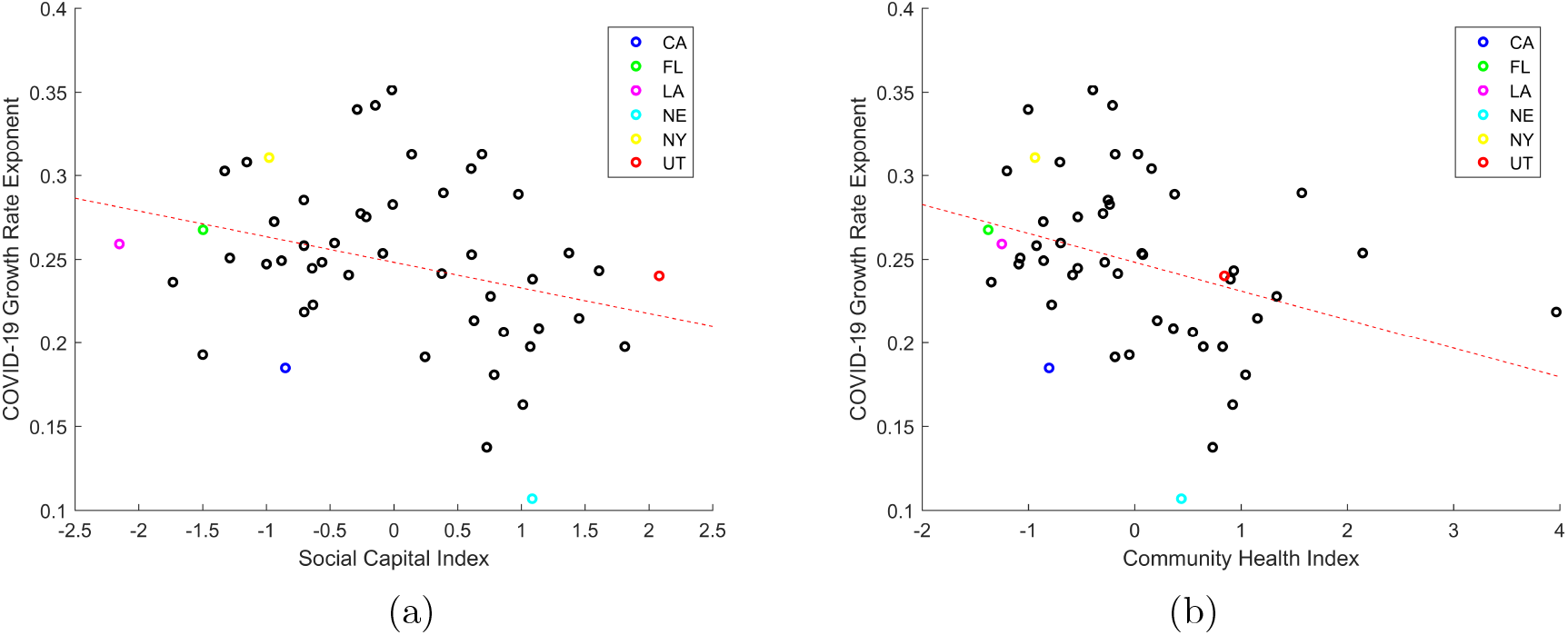
State-level fitted COVID-19 growth rate exponents are statistically significantly, negatively correlated with (a) the social capital index (*p* = 0.032, *R*^2^ = 0.092) and (b) the community health index (*p* = 0.015, *R*^2^ = 0.117). Some states are specifically marked. Case data is from the New York Times; social capital and community health indices are from [6].

**Figure 2:**
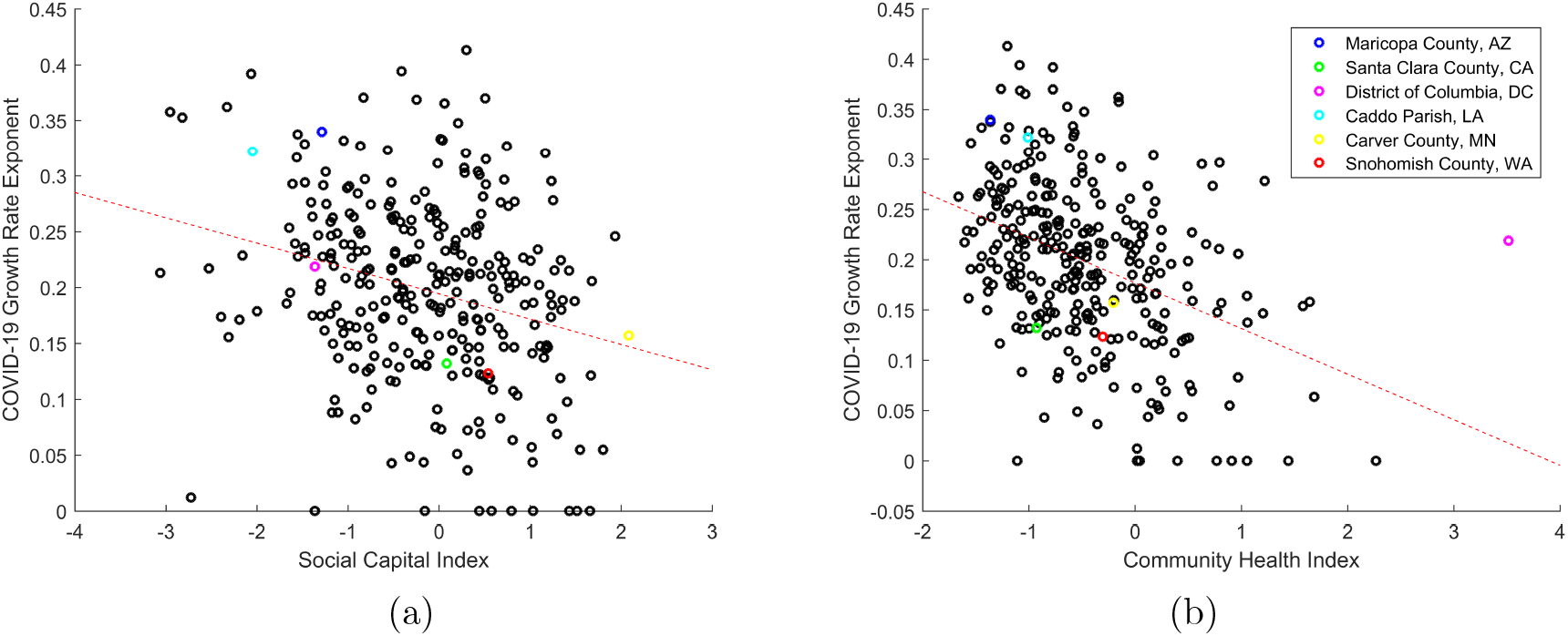
County-level fitted COVID-19 growth rate exponents are statistically significantly, negatively correlated with (a) the social capital index (*p* = 2.42 × 10^−6^, *R*^2^ = 0.0711) and (b) the community health index (*p* = 2.81 × 10^−13^, *R*^2^ = 0.162). Some counties are specifically marked (with the same colors in both panels). Case data is from the New York Times; social capital and community health indices are from [6].

Since social capital and community health are community-level attributes of local phenomena, their attributes are likely better captured at the smaller geographic level of counties than states [5]. At the county level, we find similar but stronger associations. In particular, we also find statistically significant negative correlations between fitted COVID-19 growth rate exponent and both social capital and community health indices at the county level. The community health index explains 16.2% of the variation. The effect size is such that each point of increase in social capital index corresponds to a 0.023 decrease in the growth exponent; each point of increase in the community health index corresponds to a 0.045 decrease in growth exponent. To give a sense of this impact, if the growth exponent decreased by 0.045 from 0.15 to 0.105, it cooresponds to an increase from a doubling time from 4.62 days to 6.60 days.

As a robustness check, we repeat the bivariate analysis at the county level for an alternate county-level social capital index [5] and have findings more similar to the community health index than the social capital index, see Figure 7 in Supplementary Figures, where the slope corresponds to a .041 exponent improvement for each point increase in social capital index. This corroborates a previous finding noted in [6] that the social capital index from [5] is more focused on community health factors.

As a further robustness check, we consider COVID-19 cases not just until March 31, but a further week until April 6 when fitting the growth rate exponents. These exponents may start to show greater impact from mandatory social distancing orders. As seen in Figure 8, we see similar results and in fact that social capital and community health indices now explain even more of the variation in COVID-19 growth exponents. For example, at the county-level, community health explains 20.3% of the variation.

One might wonder if our findings are largely just the result of scaling with population, in that COVID-19 growth rate exponents are greater in larger cities [18], since one might imagine there is less social capital in larger cities. Previous studies, however, show that prosocial behaviors that are included in social capital indices essentially do not scale with city size [22]. In our analysis, we find that social capital and community health indices do weakly decrease with state and county populations, but not enough to fully explain our social capital findings. For example, at the county level, the association between population and the social capital index has only (*R*^2^ = 0.023), and for the community health index has only (*R*^2^ = 0.042).

Another potential confound is that the rate of COVID-19 testing in different states and counties has been inconsistent. As shown in Fig. 3 at the state level, we find no statistically significant relationship between per capita testing rates by March 31 and social capital or community health. Even if the weak positive correlations were stronger—i.e. states with stronger social capital had more testing and more possible diagnosed cases—this would be a countervailing factor rather than an explanatory factor for our main finding of social capital being associated with reduced COVID-19 growth rate.

**Figure 3:**
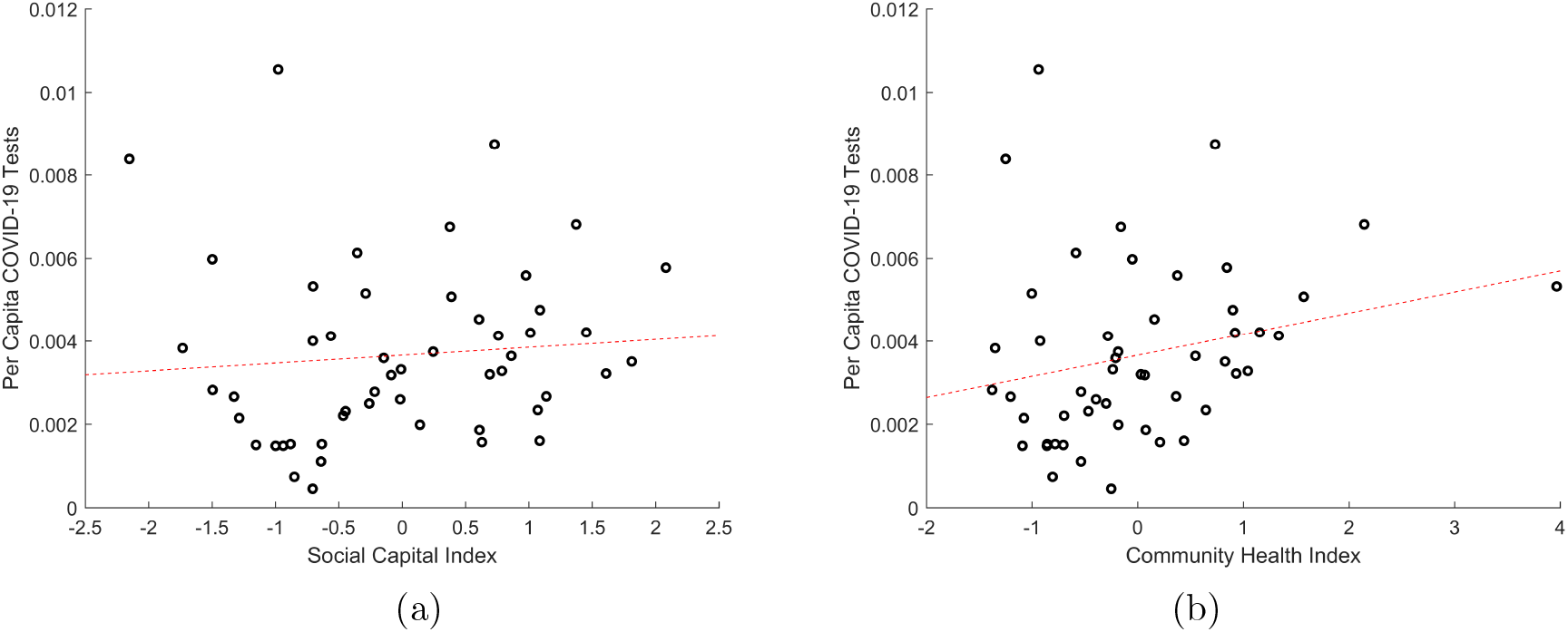
Cumulative per capita number of COVID-19 tests in U.S. states by March 31, 2020 are (a) statistically insignificantly (*p* = 0.53), very weakly positively correlated with their social capital index (*R*^2^ = 0.008) and (b) statistically insignificantly (*p* = 0.088), weakly positively correlated with their community health index (*R*^2^ = 0.058). Test count from the COVID Tracking Project; population is July 2019 estimate from the U.S. Census; social capital and community health indices are from [6].

To delve into a potential cause of social capital being associated with reduced COVID-19 growth rates and also to give some indication of whether social distancing may be more adhered to in communities with greater social capital, we consider the social capital index together with the institutional health index (which we hypothesize would be associated with following public health directives). Fig. 4 shows that the social capital index at the county level only explains a small amount of variation in reducing a coarse-grained index of mobility reduction for the week of March 23. As such, we infer that social capital is associated with reduced COVID-19 growth rate not due to coarse-grained changes in mobility but other potential causal factors.

**Figure 4:**
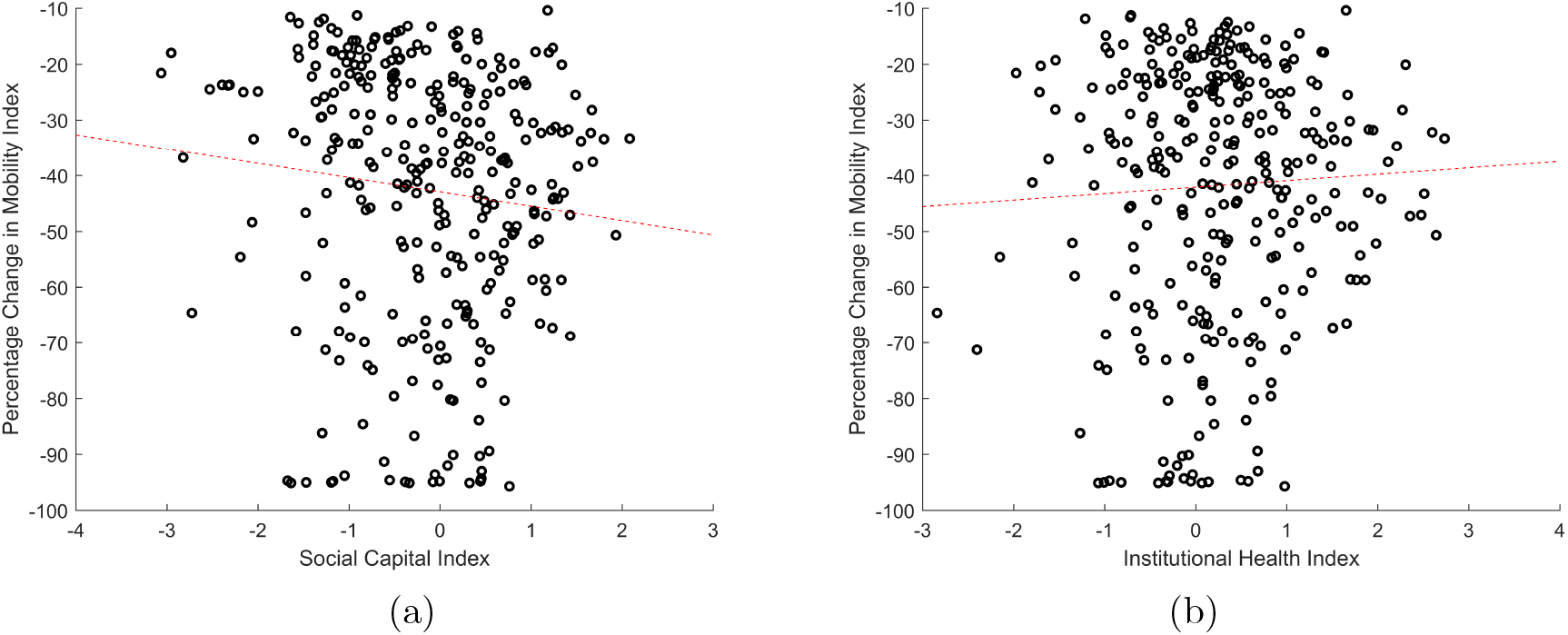
More reduction in weekly average mobility index as compared to year average in U.S. counties [restricted to those in Figure 2] for the week of March 23, 2020 has (a) weak association with greater social capital index (*p* = 0.06, *R*^2^ = 0.011), and (b) no association with institutional health index (*p* = 0.343, *R*^2^ = 0.029). Mobility index is from Cuebiq, see [20]; social capital and institutional health indices are from [6].

**Figure 5:**
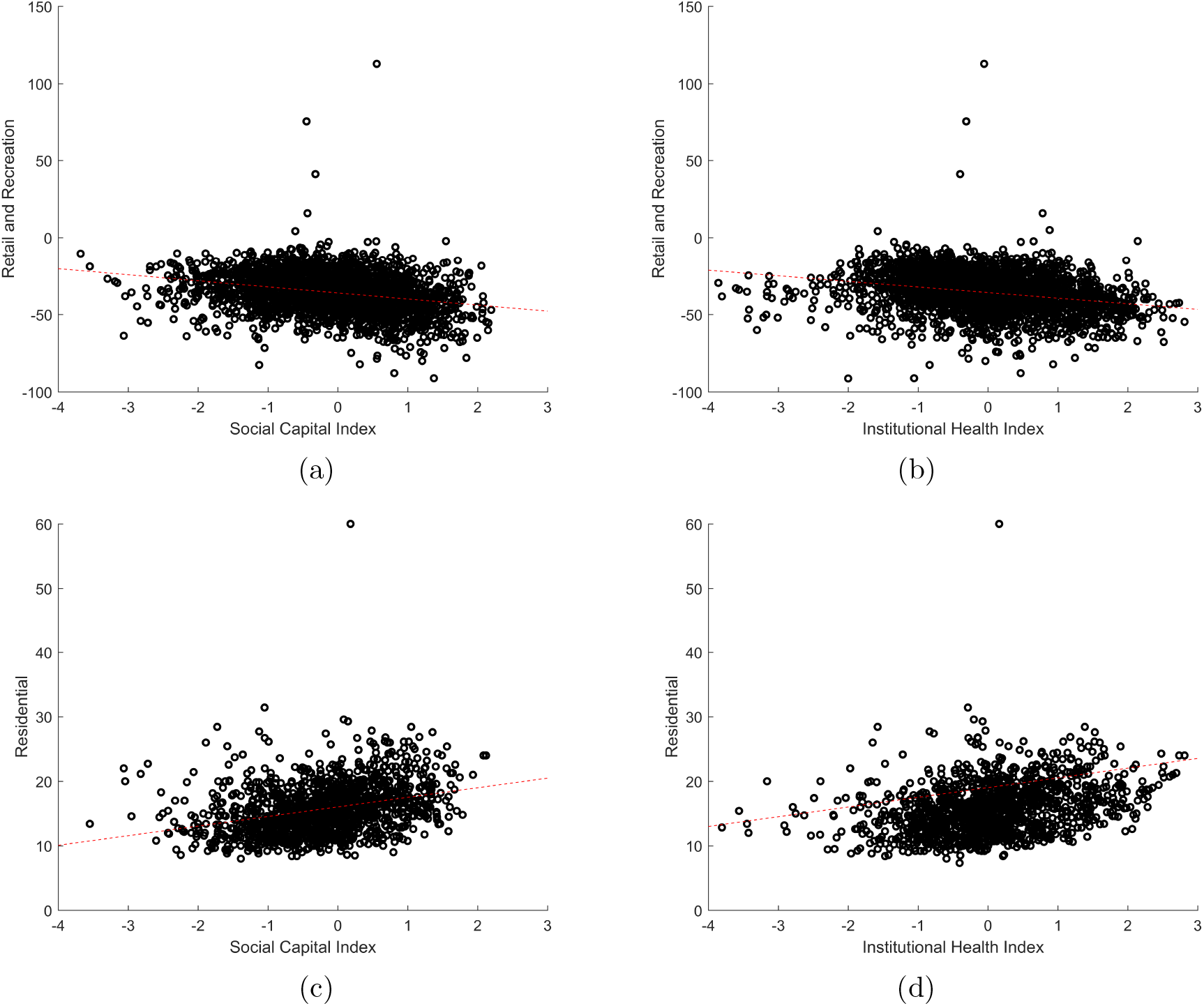
Reduced retail and recreation mobility index is associated with increased (a) social capital index (*p* = 9.13 × 10^−44^, *R*^2^ = 0.0742) and (b) institutional health index (*p* = 1.12 × 10^−41^, *R*^2^ = 0.0698). Increased residential mobility index is associated with (c) social capital index (*p* = 8.14 × 10^−33^, *R*^2^ = 0.103) and (d) institutional health index (*p* = 2.97 × 10^−30^, *R*^2^ = 0.095). Mobility indices are from Google; social capital and community health indices are from [6].

When we look at fine-grained mobility indices for the week of March 23 at the county level for nearly all counties in the United States, we see that the social capital index is associated with reduced movement in retail and recreation venues, as well as increased movement in residential venues, explaining 7.4% and 10.3% of the variation, respectively. In looking at the institutional health index, we see similar results. Not shown, but unlike for COVID-19 growth rates, social capital index is much more strongly associated than community health index. In this sense, we see that fine-grained changes in mobility (and adherence to social distancing protocols) are of interest to examine further.

## 4 Conclusion

This exploratory analysis indicates that social capital and community health are related to the growth rate of COVID-19 cases at the state and county level in the United States, one of the first such studies on the social determinants of infectious disease. We believe the observed correlations are sufficiently strong to motivate further work in this area, such as trying to tease apart causal mechanisms through experimental, quasi-experimental, or advanced statistical methods [23]. Of particular importance for causal understanding going forward is to investigate the relationship between micro-level properties of social interaction networks that will emerge from contact tracing and the area-level notions of social capital considered here. We already see that more residential movement and less retail and recreation movement are associated with area-level notions of social capital and institutional health.

Such an understanding of causal mechanisms could be used to inform potential interventions that can increase community health in communities where COVID-19 (and likely future pandemics) risk is high. Note that there is significant variation in U.S. counties with respect to susceptibility to COVID-19, not only with respect to rate of cases, but also disease severity and mortality [24]. Furthermore, it may be important to consider distinctions among communities’ associational life in planning social distancing strategies for COVID-19 contagion control in a more nuanced manner.

## Data Availability

All data is from public sources listed in the manuscript.

## 5 Supplementary Figures

**Figure 6:**
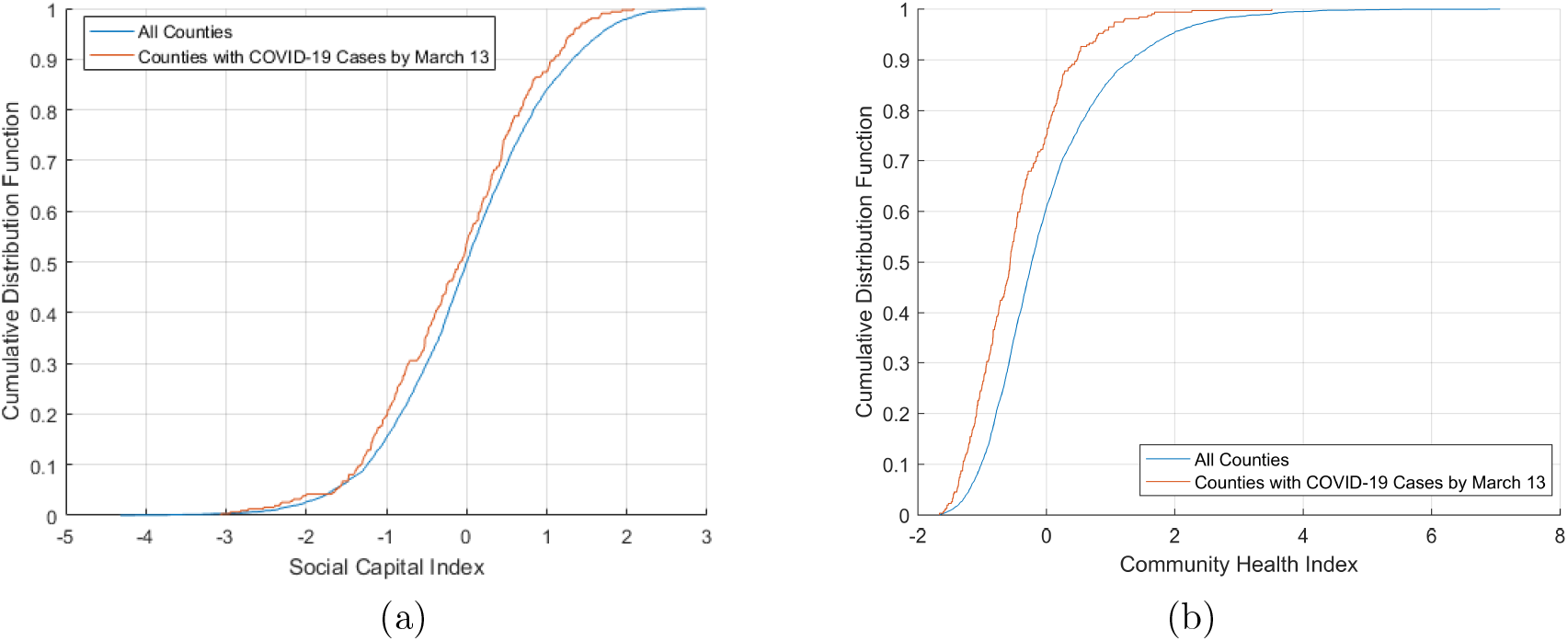
Cumulative distribution functions of (a) social capital and (b) community health indices for U.S. counties that had COVID-19 cases by March 13, 2020 are to the left of the cumulative distribution function of all counties. Case data from the New York Times; social capital and community health indices are from [6].

**Figure 7:**
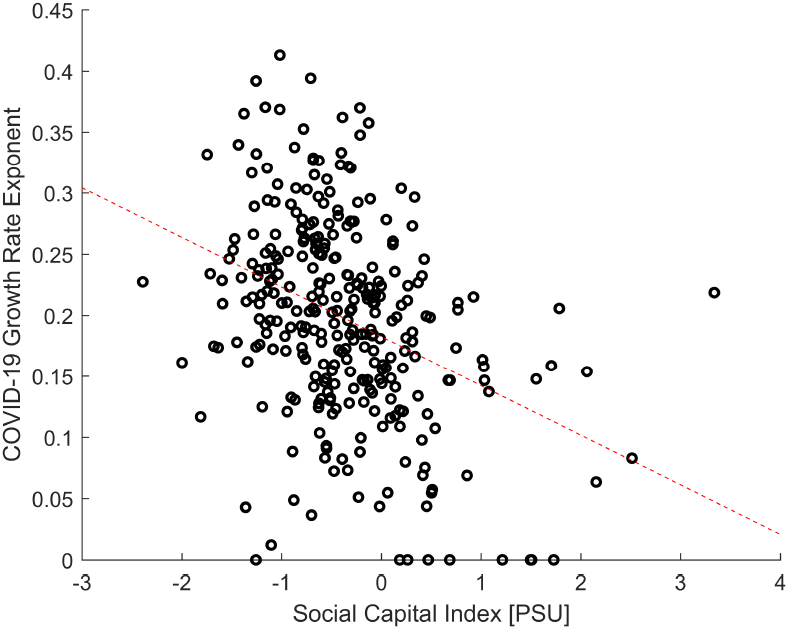
County-level fitted COVID-19 growth rate exponents are statistically significantly, negatively correlated with an alternative social capital index (*p* = 2.48 ×10^− 11^, *R*^2^ = 0.137). Case data is from the New York Times; social capital index is from [5].

**Figure 8:**
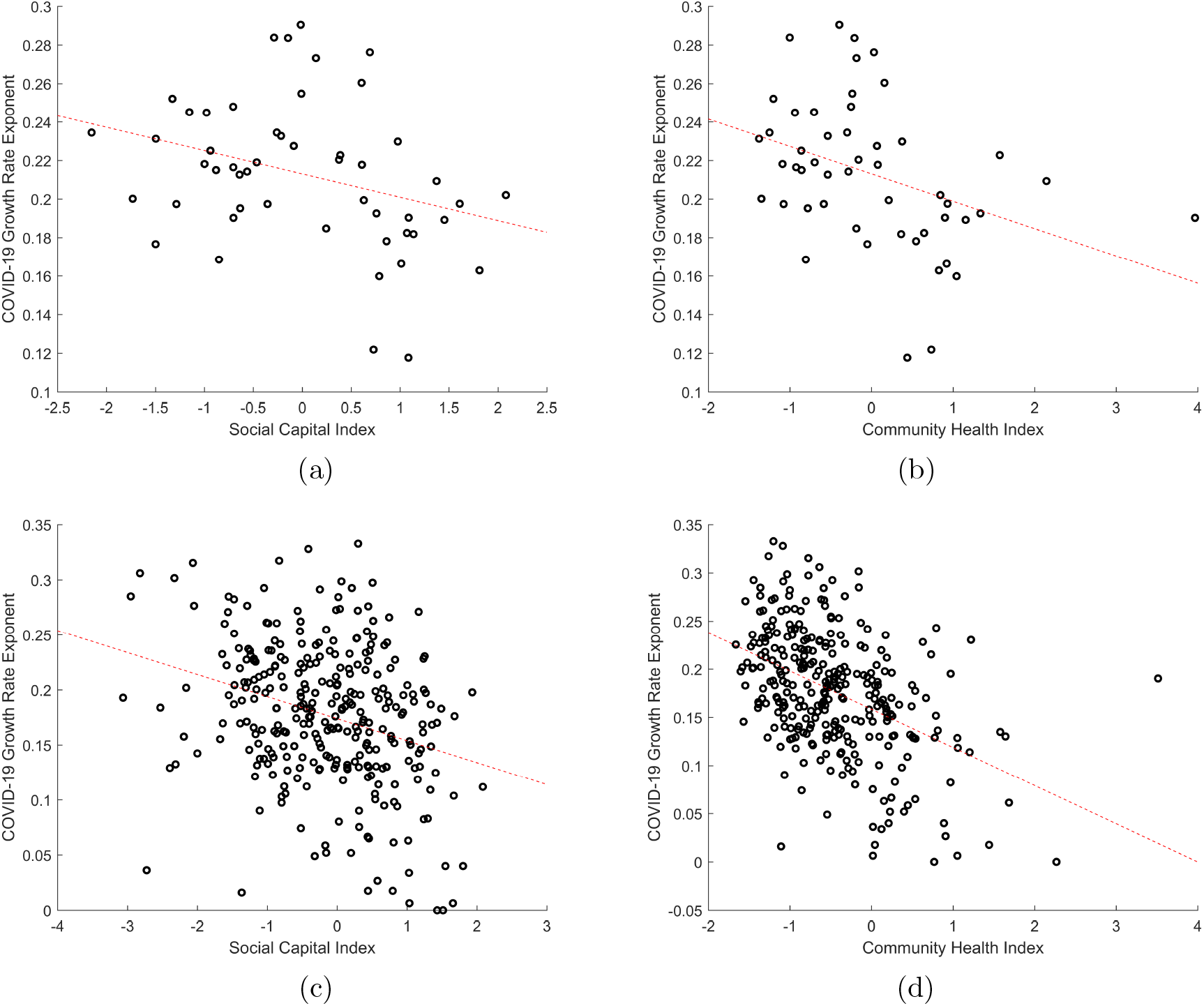
For cases upto April 6, 2020, state-level fitted COVID-19 growth rate exponents are statistically significantly, negatively correlated with (a) the social capital index (*p* = 0.023, *R*^2^ = 0.103, slope = − 0.012) and (b) the community health index (*p* = 0.0070, *R*^2^ = 0.142, slope = − 0.014). Likewise for the county-level with (c) social capital index (*p* = 9.23 × 10^− 8^, *R*^2^ = 0.0903, slope = − 0.020) (d) community health index (*p* = 1.39 × 10^− 16^, *R*^2^ = 0.203, slope = − 0.020). Case data is from the New York Times; social capital and community health indices are from [6].

## Footnotes

1 Note that Oglala Lakota County is mislabeled by its old name, Shannon County, which we corrected; and that there were ambiguities in the names of several counties in Virginia that have similar names, which we resolved by assuming the first set of data would correspond to the city. These include Alexandria City County and Alexandria County, Fairfax City County and Fairfax County, Roanoke City County and Roanoke County, and Richmond City County and Richmond County.

## Notes

### Competing Interest Statement

The authors have declared no competing interest.

### Funding Statement

No external funding was received for this work.

